# The protocol for a scoping review analyzing the association between physical activity interventions after a genitourinary cancer diagnosis and incidences of depression and anxiety

**DOI:** 10.1101/2021.07.03.21259872

**Authors:** Sydney Grob, Lorelei Mucci, Colleen McGrath

## Abstract

The following is a protocol for a planned scoping review of the effect of physical activity interventions on incidences of depression and anxiety incidences in men diagnosed with a genitourinary cancer. If exercise interventions are found to correlate with lower incidences of depression and anxiety, physical activity interventions offer a potentially modifiable, low-cost, accessible, and natural intervention to improve quality of life outcomes in men with genitourinary cancers. We plan to screen literature from publicly available databases, PubMed and Embase, for intervention studies in this area by implementing abstract/title and full-text screening by two independent reviewers followed by data extraction performed by two independent reviewers. Results will be synthesized in narrative format accompanied by relevant tabulated findings. We anticipate no ethical risks while completing this scoping review. Upon completion of this scoping review, results will be disseminated via publication.

**Article Summary (Strengths and Limitations):**

- This scoping review will address a critical area of potentially-modifiable interventions to improve quality-of-life indicators in genitourinary cancer survivorship research.
- This scoping review could inform the use of readily available, low-cost, and low-barrier, treatment options to men suffering from depression and anxiety.
- This review is limited to information from interventional studies only as a preliminary review of available research.
- This review is limited to interventional studies in male cancer patients of specific genitourinary cancers (prostate, bladder, kidney, testicular).

## Introduction

Genitourinary cancers present a major public health burden. The United States estimated 440,864 bladder, 1,414,259 prostate, 74,458 testicular, 36,068 penile, and 271,249 kidney cancer diagnoses in men during 2020[1]. In a recent cross-sectional analysis, results indicated that 5.6% (95% Confidence Interval: 4.5-6.7) of individuals diagnosed with a genitourinary cancer suffered from depression[2]. More broadly, studies have suggested that 47% of cancer patients have clinically apparent psychiatric disorders, with most being anxiety and depression, and 90% of these disorders were attributed to disease and disease treatment[3]. Given the high prevalence of poor mental health among cancer patients, it is important to understand how to mitigate the severity of anxiety and depression in this population to improve quality of life outcomes for these patients.

A cancer diagnosis has been associated with poor mental health outcomes including cancer-related fatigue, cognitive impairment, sleep problems, depression, pain, anxiety, and physical dysfunction[4]. Many studies have described the benefit of exercise interventions in improving psychological outcomes in a myriad of populations[5-7]. More specifically, many studies have evaluated the potential relationship between various psychological outcomes in many cancer types including lung[8, 9], breast[10, 11], prostate[12, 13], bladder[14], kidney[15], testicular [16], and additional cancers[17]. For example, in a study evaluating walking interventions in patients with lung cancer, results indicated that the introduction of an exercise intervention improved anxiety (P=0.009 and P=0.006 three and six months at follow-up) and depression (P=0.00006 and 0.004 three and six months at follow-up) levels over time[8]. Furthermore, another study found that high intensity interval training regimens did not improve depression (P=0.81), anxiety (P=0.19), stress (P=0.22), and sleep quality (P=0.15), but it did improve cancer-related fatigue (P=0.003) and self-esteem (P=0.029) in men diagnosed with testicular cancer compared to the control group[16]. A larger synthesis of the current literature on the effect of exercise interventions on incidences of depression and anxiety in men diagnosed with a genitourinary cancer is necessary.

The primary aim of this study is to summarize the current literature’s evidence surrounding the association of exercise interventions and incidences of depression and anxiety in men diagnosed with genitourinary cancers. We are specifically interested in this question because exercise interventions, if found to be correlated with decreasing depression and anxiety incidences, offer a potentially modifiable, low-cost, accessible, and natural intervention to improve quality of life outcomes in men with genitourinary cancers.

### Review Questions

1. Is there an association between physical activity interventions after a genitourinary cancer diagnosis and incidences of depression and anxiety?

## Methods and Analysis

### Searches

We will leverage PubMed and EMBASE databases. We will only include interventional studies, but there will be no parameters placed on publication date, language, or journal. We detail full search strategy below and include a list of search terms in the appendix.

### Search Strategy

#### Types of studies to be included

Only interventional studies/materials will be included in this scoping review.

#### Condition or domain being studied

Psychosocial outcomes (depression, anxiety) Search parameters for the genitourinary cancers will be employed, including terms such as, but not limited to, “cancer,” “neoplasm,” “malignancy,” “tumor,” “carcinoma,” coupled with terms such as, but not limited to, “testicle,” “testes,” “scrotal,” “genital,” “prostate,” “bladder,” “kidney,” “renal,” and “urinary.” Search parameters for exercise interventions included terms for the notion of intervention itself (“intervention,” “trial,” “program”) paired with specific exercises, such as but not limited to the following terms, “strength,” “running,” “walking,” “stairs,” “cycling,” “spinning,” “weight,” and more. Search parameters for the psychosocial outcomes of interest included “depression,” “depressive,” “anxiety,” and “anxious.” A full search strategy delineated for each database can be provided in separate documentation.

### Participants/Population

We are interested in studying men of all ages who received a clinical diagnosis of bladder cancer, kidney cancer, prostate cancer and testicular cancers followed by enrollment in an exercise intervention following diagnosis. We will exclude murine or other animal models of any genitourinary cancer.

### Intervention(s), exposure(s)

Our exposure of interest will include any exercise intervention that has been hypothesized and/or shown to improve psychological, physical, or overall survival in men diagnosed with a genitourinary cancer diagnosis. Below are some, but not all, of the exercise interventions of interest:

Cycling/biking/spinning, running, walking/hiking, swimming, calisthenics, rowing, stair master, ski machine, yoga, squash, racquetball, tennis, hiking, weight training, high intensity interval training, resistance training, jogging

### Comparator(s)/control

Men diagnosed with a genitourinary cancer who did not undergo an exercise intervention

#### Inclusion Criteria

- Studies that include diagnosed genitourinary cancer patients (all ages)
- Studies that include men diagnosed with genitourinary cancers
- Studies that include an exercise intervention in diagnosed genitourinary cancer patients
- Studies that report measure(s) of association (i.e. HR, OR, RR) for depression and/or anxiety
- Studies that quantitatively measure or assess depression and anxiety

#### Exclusion Criteria

- Studies that are not interventional (i.e. observational, case studies, reviews, book chapters, editorials, correspondence)
- Interventional studies not conducted in humans
- Studies which do not separate male and female outcome measurements for applicable genitourinary cancers (i.e. bladder, kidney)
- Studies focused solely on any cancer except bladder, kidney, testicular, prostate
- Studies which only include female participants
- Studies that do not quantitatively measure or assess depression and anxiety

### Main Outcome(s)

Research Question 1: depression, anxiety (relative risk, odds ratio, hazard ratio, rate ratio)

### Data extraction (selection and coding)

- Selection: Independent screening by two reviewers for inclusion and exclusion criteria. Instances where independent reviewers do not agree as to whether to include or exclude an article will be resolved in the following ways: 1) reach consensus by discussion; 2) if consensus is not reached by discussion, have a third independent reviewer make a determination.
  - Primary screening will involve both readers independently reading abstracts and deciding whether to include or exclude article based on laid out criteria.
  - Secondary screening will involve reading the entire article and documenting why articles have been excluded at this stage in a PRISMA-compliant flow diagram.
- Coding: For included articles following the second screening, the below information will be extracted and documented for each article:
  - Summary information for each article:
  - Details of Interventional Study:
  - Study Population/Demographics:
  - Genitourinary Cancer Studied:
  - Details of Exercise Intervention:
  - Details of Measurement of Outcome Variable:
  - Results: Measure of association between exercise intervention and outcome:
  - Additional results as applicable
  - Study Strengths and Limitations and relevant discussion points:

### Strategy for data synthesis

We will perform a description synthesis of our findings. Supporting tables and figures will complement this synthesis to provide more robust information on included studies and key findings.

### Analysis of subgroups of subsets

We will study all genitourinary cancers together. We will also conduct a subset analysis by genitourinary cancer type.

## Data Availability

We plan to screen literature from publicly available databases including PubMed and Embase. We will synthesize our findings in a narrative format and aim to publish in a publicly available journal.

## Contact details for further information

Sydney Grob

Colleen McGrath

## Organizational affiliation of the review

Harvard T.H. Chan School of Public Health https://www.hsph.harvard.edu/

### Ethics and Dissemination

(ethical and safety considerations and any dissemination plan)

There are no risks related to the conduct of this scoping review. Upon completion of the review, we plan to disseminate our findings through publication.

## Authors’ Contributions

This scoping review will be conducted by the following:

Sydney Grob, Harvard T.H. Chan School of Public Health

Lorelei Mucci, Harvard T.H. Chan School of Public Health

Colleen McGrath, Harvard T.H. Chan School of Public Health

Authors S.G. and C.M. will conduct the screening, extraction, data synthesis, and manuscript composition. Author L.M. will be available for consensus in screening and extraction processes as needed as well as contribute to manuscript composition.

Anticipated or actual start date: 05/01/2021

Anticipated completion date: 07/31/2021

